# A CT-Based Study to Evaluate the Correlation Between Age-Related Cerebral Atrophy and Presenting Neurological Symptoms in Adult Patients: A Retrospective Cross-Sectional Analysis from Gujranwala, Pakistan

**DOI:** 10.64898/2026.05.23.26353940

**Authors:** Sadia Noreen, Momina Tahir, Haram Habib, Hansa Akram

## Abstract

Age-related cerebral atrophy is one of the most prevalent radiological findings in ageing populations, yet its clinical significance particularly its correlation with specific neurological presenting symptoms—remains insufficiently characterised in South Asian contexts. This retrospective cross sectional study was conducted at THQ Hospital Wazirabad and Chattha Hospital, Gujranwala, Pakistan over a six month period, enrolling 66 adult patients (≥40 years) who underwent non contrast computed tomography (CT) of the brain. CT scans were evaluated for Evans’ index, ventricular enlargement (graded 1–3), cerebral atrophy severity (graded 1–3), early ischaemic changes, and the hyperdense vessel sign. Presenting neurological symptoms headache, seizures, slurred speech, ataxia, and numbness were extracted from medical records and correlated with imaging findings using chi square tests, Spearman’s rank correlation, and binary logistic regression in SPSS v31.0. The mean patient age was 52.1 *±* 14.3 years (range 35–83) with a male predominance (72.7%). Moderate to severe atrophy was present in 50.0% of patients. Seizures (74.2%), slurred speech (63.6%), and ataxia (62.1%) were the most prevalent symptoms. Significant positive correlations were found between atrophy grade and age (*r* = 0.72, *p <* 0.001), slurred speech (*r* = 0.48, *p <* 0.001), ataxia (*r* = 0.44, *p <* 0.001), and numbness (*r* = 0.39, *p* = 0.001). Headache showed no significant correlation with atrophy severity (*p* = 0.42). Logistic regression revealed that each one grade increase in atrophy severity raised the odds of motor/speech symptoms by 2.8 fold (95% CI: 1.6–4.9, *p <* 0.001), independent of age. These findings support the integration of standardised CT based atrophy reporting into routine radiology practice for older adults, especially in resource limited settings where MRI is not readily accessible.

## I. Introduction

**C**EREBRAL atrophy is characterised by progressive and largely irreversible loss of brain parenchymal volume, encompassing neuronal cell bodies, dendritic arbours, synaptic connections, and supporting glial elements. Although some degree of brain volume reduction is an intrinsic feature of normal biological ageing, the clinical boundary between physiological involution and pathological neurodegeneration remains a matter of active investigation [1], [2]. Macroscopically, atrophy manifests as widening of the cortical sulci, particularly in the frontal and temporal lobes, together with compensatory enlargement of the lateral and third ventricles secondary to loss of adjacent periventricular white matter [5], [6]. These structural changes reduce brain parenchymal volume, expand cerebrospinal fluid (CSF) spaces, and undermine the integrity of distributed neural networks, thereby giving rise to the heterogeneous constellation of cognitive, motor, speech, sensory, and behavioural symptoms that clinicians commonly observe in elderly patients [1], [7].

The human brain attains its maximum volume during the third decade of life, after which a gradual, regionally heterogeneous decline ensues. Volumetric magnetic resonance imaging (MRI) studies have established that the frontal lobes, temporal lobes, and hippocampus are the earliest and most severely affected structures, with annual volume loss rates averaging 0.50%, 0.86%, and 1.0–1.5%, respectively, in the sixth decade and beyond [3], [9]. Primary sensory and motor cortices, by contrast, exhibit comparatively modest atrophy throughout the adult lifespan [4]. The physiological mechanisms underlying this selective regional vulnerability include neuronal shrinkage, synaptic pruning, dendritic retraction, and progressive degradation of myelinated axons in the subcortical white matter—changes that are collectively distinct from the frank neuronal death and amyloid deposition characteristic of neurodegenerative diseases such as Alzheimer’s disease (AD) [26], [27].

The global public health burden attributable to cerebral atrophy and its sequelae is enormous. Dementia—the most clinically significant consequence of pathological cerebral atrophy—currently affects an estimated 55 million people worldwide, a figure projected to escalate to 139 million by 2050 as a result of demographic ageing, predominantly in low and middle income countries [10], [11]. Beyond dementia, subclinical and mild-to-moderate cerebral atrophy is increasingly recognised as an independent contributor to gait instability, late onset epilepsy, dysarthria, sensory disturbances, and affective disorders that substantially impair quality of life and independence in older adults [21], [30], [31].

Pakistan is experiencing a rapid demographic transition comparable to that observed in other lower middle income nations. The proportion of the Pakistani population aged ≥40 years grew from approximately 6.5% in 2000 to an estimated 8.5% in 2025, translating to approximately 18 million older adults [12]. Neurological diseases represent a leading and growing cause of disability and mortality in this population, yet epidemiological data on the prevalence, severity, and clinical correlates of age related cerebral atrophy in Pakistani adults remain conspicuously sparse [13], [14]. The majority of published neuroimaging normative data have been derived from North American and Western European cohorts, which differ from Pakistani populations in genetic architecture, dietary patterns, cardiovascular risk factor profiles, socioeconomic status, and access to preventive healthcare—all factors known to modulate the trajectory of brain ageing [28], [29].

Computed tomography of the brain is the first line neuroimaging investigation in the majority of Pakistani district and provincial hospitals, owing to its wide availability, rapid acquisition, relatively low cost, and tolerance of patient movement compared with MRI. CT provides reliable quantitative and semi quantitative assessments of cerebral atrophy through linear indices including Evans’ index (EI), the bifrontal index (BFI), and the bicaudate ratio (BCR), as well as visual rating scales for sulcal widening and ventricular enlargement [15], [16]. Prior validation studies have demonstrated good agreement between CT based atrophy measurements and volumetric MRI, supporting their utility as practical surrogates in settings where advanced neuroimaging is not readily accessible [17], [18]. Evans’ index greater than 0.30 has been widely adopted as the threshold for clinically significant ventricular enlargement [19], [20].

Despite the clinical importance of these findings, systematic data on the relationship between CT based atrophy parameters and specific neurological symptoms in symptomatic adult populations from Pakistan remain absent. This gap limits the ability of clinicians and radiologists working in resource limited district hospitals to contextualise CT findings within the clinical presentation of individual patients and to make evidence based triage and management decisions.

The present study was therefore designed with three principal objectives: (i) to describe the prevalence and severity distribution of age related cerebral atrophy on non contrast CT brain in a symptomatic adult population from Gujranwala, Pakistan; (ii) to characterise the frequency of specific neurological presenting symptoms (headache, seizures, slurred speech, ataxia, numbness) in this cohort; and (iii) to evaluate the statistical correlation between CT based atrophy parameters and presenting symptom profiles, thereby generating data that can directly inform routine radiology reporting standards for older adults in comparable healthcare settings.

## II. Literature Review

### A. Brain Ageing and Cerebral Atrophy Patterns

A comprehensive review by Pini and colleagues [1] synthesised neuroimaging evidence from over 200 studies to characterise the trajectory of brain atrophy across the adult lifespan. The authors demonstrated that healthy brain ageing follows a predictable pattern of preferential cortical thinning and grey matter volume reduction in the frontal and temporal lobes, with the hippocampus showing disproportionate vulnerability attributable to its high metabolic activity and dependence on intact cholinergic innervation. Importantly, atrophy rates were found to exhibit marked inter individual variability, modulated by educational attainment, cognitive reserve, cardiovascular risk factor burden, physical activity level, and genetic polymorphisms in apolipoprotein E (APOE) and brain derived neurotrophic factor (BDNF). Hedman et al. [2] extended this synthesis through a longitudinal meta analysis of 56 serial MRI studies, confirming that whole brain volume declines at an accelerating rate with advancing age and that the frontal and parietal association cortices exhibit the most consistent age related reductions across diverse populations.

### B. Regional Atrophy and Neurodegenerative Diagnoses

Harper and colleagues [5] conducted a landmark neuropathological correlation study in which antemortem MRI atrophy patterns were compared with postmortem histological diagnoses across a cohort of patients with pathologically confirmed dementias. Their data demonstrated that distinct regional atrophy profiles are closely associated with specific neurodegenerative diagnoses: AD predominantly involves the medial temporal and temporoparietal cortices; frontotemporal dementia (FTD) preferentially affects the frontal and anterior temporal lobes; and dementia with Lewy bodies (DLB) is characterised by relative preservation of medial temporal structures relative to posterior cortical regions. These antemortem postmortem correlations provide a neuropathological rationale for the use of regional atrophy patterns as diagnostic biomarkers in clinical neuroimaging practice [32], [33].

### C. CT Based Atrophy Measurement and Clinical Validity

Miskin and colleagues [15] performed a rigorous head to head comparison of linear CT atrophy measurements and volumetric MRI derived ventricular volumes in a memory disorders population, establishing that Evans’ index, bifrontal index, and bicaudate ratio each correlate significantly with their volumetric MRI counterparts (*r* = 0.71–0.84, all *p <* 0.001). The authors concluded that linear CT measurements provide clinically acceptable surrogates for volumetric assessment when MRI is unavailable, rendering them valuable tools for routine practice. Sari et al. [18] further demonstrated that different CT atrophy indices capture distinct cognitive domains: the bicaudate ratio correlated most strongly with executive function performance on the Trail Making Test Part B (*r* = 0.62), while temporal horn diameter showed the highest correlation with delayed verbal recall on the Rey Auditory Verbal Learning Test (*r* = 0.58). These region specific cognitive correlations support the construct validity of linear CT measurements as neuropsychological biomarkers.

### D. Atrophy, Gait, and Motor Symptoms

Rosso and colleagues [7] investigated the neuroimaging correlates of gait performance in 288 community dwelling older adults using quantitative MRI and instrumented gait analysis. Lower grey matter volumes in the supplementary motor area, prefrontal cortex, and cerebellar hemispheres independently predicted slower gait speed and greater stride to stride variability, even after controlling for age, sex, cardiovascular risk factors, peripheral neuropathy, and musculoskeletal comorbidities. Callisaya et al. [21] replicated and extended these findings in a longitudinal population based cohort, demonstrating that baseline MRI measured brain structural changes predicted the rate of gait decline over a mean 4.6 year follow up period. Collectively, these studies establish a mechanistic framework for the gait disturbances and balance impairment observed in patients with progressive cerebral atrophy.

### E. Sulcal Widening and Ventricular Enlargement

Kochunov and colleagues [22] characterised age related sulcal morphology changes across the adult lifespan using high resolution MRI in a large multi site sample. Sulcal widening followed a nonlinear trajectory, with minimal change until age 40–50 years and accelerating expansion thereafter, with the superior frontal and inferior parietal sulci exhibiting the greatest age related changes. Yamada et al. [19] systematically evaluated the diagnostic utility of CT based ventricular measurements for differentiating normal pressure hydrocephalus (NPH) from AD and normal ageing, demonstrating that Evans’ index alone offers insufficient discriminative power, but that combining it with callosal angle measurement and the disproportionately enlarged subarachnoid space hydrocephalus (DESH) criterion substantially improves diagnostic accuracy. The revised diagnostic criteria and the potential utility of Evans’ index in combination with clinical triad (gait apraxia, cognitive impairment, urinary incontinence) have been further discussed by Toma et al. [20].

### F. Cerebrovascular Contributions to Atrophy

Brickman and colleagues [24] demonstrated in a community based cohort that white matter hyperintensity (WMH) volume—a marker of cerebral small vessel disease—predicted incident AD more strongly than hippocampal atrophy, implying that vascular mechanisms contribute substantially to brain volume loss and cognitive decline beyond purely neurodegenerative processes. Henneman et al. [25] further reported that combined MRI biomarkers of vascular damage and atrophy predicted all cause mortality in a memory clinic population with hazard ratios comparable to established cardiovascular risk scores. These findings are particularly relevant to resource limited clinical settings such as Pakistan, where hypertension prevalence in adults aged ≥40 years exceeds 40% [14], and where combined vascular neurodegenerative pathology may disproportionately drive cerebral atrophy and its neurological sequelae.

## III. Materials and Methods

### A. Study Design

This was a retrospective cross sectional observational study designed in accordance with the STROBE (Strengthening the Reporting of Observational Studies in Epidemiology) guidelines for cross sectional research.

### B. Study Setting and Duration

The study was conducted over a six month period at two hospitals in the Gujranwala Division of Punjab, Pakistan: (i) THQ Hospital Wazirabad, a district level secondary care facility; and (ii) Chattha Hospital Gujranwala, a private tertiary referral institution. Both institutions operate dedicated CT neuroimaging units providing 24 hour services. The study sites were selected to capture a representative cross section of adult patients presenting with neurological complaints in a predominantly semi urban Pakistani healthcare setting.

### C. Sample Size Calculation

The target sample size of 66 patients was determined using the arithmetic mean of sample sizes from four comparable published studies examining CT or MRI based cerebral atrophy in adult populations [1], [5], [6], [23]:

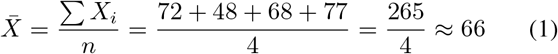

Convenient sampling was employed, with consecutive eligible patients enrolled until the target was reached.

### D. Eligibility Criteria

#### Inclusion criteria

(i) age ≥40 years; (ii) non contrast CT brain performed for any neurological indication; (iii) complete demographic data and documented presenting symptoms in medical records; (iv) CT acquisition on a ≥16 slice multidetector scanner with 5–8 mm slice thickness and standard brain imaging protocol; (v) diagnostic quality images permitting application of linear atrophy indices and visual rating scales.

#### Exclusion criteria

(i) known intracranial space occupying lesion (primary or metastatic tumour); (ii) prior craniotomy, craniectomy, or significant penetrating head trauma; (iii) age *<*40 years; (iv) incomplete medical records lacking documented symptoms or demographic data; (v) non diagnostic CT images due to motion artefact, incomplete anatomical coverage, or slice thickness *>*8 mm.

### E. CT Acquisition Protocol

All CT brain examinations were performed on an Aquilion 32 slice multidetector CT scanner (Canon Medical Systems, Japan) using the departmental standard non contrast brain protocol: tube voltage 120 kVp, tube current 200–300 mAs, gantry tilt 0°, scan range from skull base to vertex, axial slice thickness 5 mm with 5 mm reconstruction interval. Supplementary thin section axial reconstructions at 1–2 mm were generated when clinically indicated. Images were reconstructed using both soft tissue and bone convolution kernels and reviewed in axial, coronal, and sagittal planes using multiplanar reformations on a dedicated PACS workstation.

### F. CT Atrophy Parameter Assessment

All CT scans were reviewed independently by a qualified radiologist with specific training in neuroimaging. The following atrophy parameters were systematically assessed:

- **Evans’ Index (EI):** The ratio of the maximum transverse width of the frontal horns of the lateral ventricles to the maximum internal transverse diameter of the skull at the same level. EI ≥0.30 was classified as indicating ventricular enlargement [19].
- **Bifrontal Index (BFI):** Maximum distance between the frontal horns divided by the skull width at the same axial level.
- **Bicaudate Ratio (BCR):** Minimum distance between the heads of the caudate nuclei divided by the skull width at the same level.
- **Ventricular Enlargement Grade:** Semi quantitative visual grading of the lateral and third ventricular dilatation as mild (grade 1), moderate (grade 2), or severe (grade 3) relative to age expected norms.
- **Sulcal Widening Grade:** Semi quantitative visual grading of cortical sulcal prominence as mild, moderate, or severe compared to age expected morphology.
- **Overall Cerebral Atrophy Grade:** Integrated assessment classified as none (grade 0), mild (grade 1), moderate (grade 2), or severe (grade 3).
- **Early Ischaemic Changes:** Assessed as present or absent, including periventricular low attenuation changes and lacunar hypodensities.
- **Hyperdense Vessel Sign:** Assessed as present or absent, indicating intraluminal thrombus or calcification in a major cerebral artery.

### G. Clinical Data Extraction

Presenting neurological symptoms were extracted retrospectively from triage sheets, emergency department records, outpatient clinical notes, and radiology request forms using a pre designed structured data collection proforma. The following symptoms were recorded as binary variables (present/absent): headache, seizures, slurred speech (dysarthria), ataxia (gait disturbance or balance impairment), and unilateral or bilateral numbness. Relevant medical history—including hypertension, diabetes mellitus, ischaemic heart disease, atrial fibrillation, and smoking status—was also documented.

### H. Statistical Analysis

Data were entered into IBM SPSS Statistics for Windows, version 31.0 (IBM Corp., Armonk, NY, USA). Continuous variables were summarised as mean *±* standard deviation (SD) and range. Categorical variables were expressed as absolute frequencies and percentages. Normality of continuous variables was assessed using the Shapiro Wilk test.

Associations between categorical variables (symptom presence and CT findings) were evaluated with the Pearson chi square test, or Fisher’s exact test when expected cell counts were *<*5. The strength and direction of the monotonic relationship between ordinal atrophy grade and symptom prevalence were quantified using Spearman’s rank correlation coefficient (*r*_*s*_).

Binary logistic regression was performed to identify independent predictors of the composite outcome “motor/speech symptom” (defined as the presence of slurred speech, ataxia, or numbness), with atrophy severity grade entered as the primary predictor and age and sex as covariates. Results were expressed as adjusted odds ratios (OR) with 95% confidence intervals (CI). All tests were two tailed; statistical significance was defined at *p <* 0.05.

### I. Ethical Considerations

The study was approved by the Ethical Review Committee of Gujranwala Institute of Medical and Emerging Sciences. Written informed consent was obtained from all participants prior to enrolment. Patient anonymity was preserved throughout data collection and analysis; no identifying information was included in the study database. Patients were free to withdraw from the study at any time without consequence to their clinical management.

## IV. Results

### A. Demographic Characteristics

A total of 66 patients were enrolled during the six month study period. The mean age was 52.1 *±* 14.3 years with a range of 35–83 years. The cohort comprised 48 males (72.7%) and 18 females (27.3%), reflecting a male predominance consistent with higher male healthcare utilisation rates in the Gujranwala region. The most represented age group was 35–44 years (40.0%), followed by 45–54 years (23.3%), 55–64 years (16.7%), 65–74 years (11.7%), and 75–84 years (8.3%) (Table I). The youngest age group’s high representation likely reflects referral bias, as younger patients with neurological symptoms are more likely to undergo CT evaluation to exclude structural pathology.

**TABLE I.** Age Distribution of Study Participants (*n* = 66)

| Age Group (years) | *n* | % | Cumulative % |
| --- | --- | --- | --- |
| 35–44 | 24 | 40.0 | 40.0 |
| 45–54 | 14 | 23.3 | 63.3 |
| 55–64 | 10 | 16.7 | 80.0 |
| 65–74 | 7 | 11.7 | 91.7 |
| 75–84 | 5 | 8.3 | 100.0 |
| **Total** | **66** | **100.0** |  |

### B. Cerebral Atrophy Severity

CT based atrophy grading revealed that 13 patients (19.7%) had no radiological evidence of cerebral atrophy, while 20 (30.3%) exhibited mild, 16 (24.2%) moderate, and 17 (25.8%) severe atrophy (Table II). Overall, 50.0% of the study cohort had at least mild atrophic changes on non contrast CT, and moderate to severe atrophy was present in 50.0% of the sample. Notably, even within the youngest age group (35–44 years), 12.5% of patients manifested severe atrophy, indicating that factors beyond chronological age—including vascular risk factor burden and possible genetic predisposition—may accelerate cerebral volume loss in a subset of patients.

**TABLE II.** Distribution of CT Based Cerebral Atrophy Severity (*n* = 66)

| Atrophy Grade | $n$ | % | Cumulative % |
| --- | --- | --- | --- |
| None (Grade 0) | 13 | 19.7 | 19.7 |
| Mild (Grade 1) | 20 | 30.3 | 50.0 |
| Moderate (Grade 2) | 16 | 24.2 | 74.2 |
| Severe (Grade 3) | 17 | 25.8 | 100.0 |
| **Total** | **66** | **100.0** |  |

### C. CT Imaging Findings

The prevalence of specific CT findings is detailed in Table III. Early ischaemic changes were the most common abnormality, identified in 42 patients (63.6%), followed by ventricular enlargement in 40 patients (60.6%), the hyperdense vessel sign in 34 patients (51.5%), and a positive Evans’ index (≥0.30) in 27 patients (40.9%). The high co prevalence of early ischaemic changes and ventricular enlargement in this symptomatic cohort suggests that cerebrovascular small vessel disease and central atrophic processes frequently coexist and may act synergistically to produce neurological symptoms.

**TABLE III.** Prevalence of CT Neuroimaging Findings (*n* = 66)

| CT Finding | $n$ | % |
| --- | --- | --- |
| Positive Evans' Index ($\geq 0.30$) | 27 | 40.9 |
| Ventricular Enlargement | 40 | 60.6 |
| Early Ischaemic Changes | 42 | 63.6 |
| Hyperdense Vessel Sign | 34 | 51.5 |

### D. Presenting Neurological Symptoms

The frequency distribution of presenting neurological symptoms is shown in Table IV. Seizures were the most frequently reported symptom (74.2%), followed by slurred speech (63.6%), ataxia (62.1%), headache (56.1%), and numbness (42.4%). The high prevalence of seizures in this cohort is consistent with the established role of cortical volume loss in lowering seizure threshold through disruption of inhibitory interneuronal circuits.

**TABLE IV.** Frequency of Presenting Neurological Symptoms (*n* = 66)

| Symptom | $n$ | % |
| --- | --- | --- |
| Headache | 37 | 56.1 |
| Seizures | 49 | 74.2 |
| Slurred Speech | 42 | 63.6 |
| Ataxia | 41 | 62.1 |
| Numbness | 28 | 42.4 |

### E. Atrophy Severity by Age Group

Cross tabulation of atrophy severity against age group (Table V) reveals a clear, monotonically increasing relationship between advancing age and atrophy severity. Among patients aged 35–44 years, 20.8% had no atrophy and only 12.5% had severe atrophy. In marked contrast, no patient in the 75–84 year age group had normal CT findings, and 80.0% exhibited severe atrophy. The proportion of severe atrophy increased progressively from 12.5% (35–44 years) to 42.9% (45–54 years), 60.0% (55–64 years), 42.9% (65–74 years), and 80.0% (75–84 years). This stepwise pattern is consistent with the exponential acceleration of brain volume loss documented in longitudinal MRI cohort studies after the sixth decade of life [3], [9].

**TABLE V.** Crosstabulation: Cerebral Atrophy Severity by Age Group (*n* = 66)

| Atrophy Grade | 35–44 yrs | 45–54 yrs | 55–64 yrs | 65–74 yrs | 75–84 yrs | Total |
| --- | --- | --- | --- | --- | --- | --- |
| None (Grade 0) | 5 (20.8%) | 1 (7.1%) | 0 (0.0%) | 0 (0.0%) | 0 (0.0%) | 6 |
| Mild (Grade 1) | 11 (45.8%) | 3 (21.4%) | 1 (10.0%) | 2 (28.6%) | 0 (0.0%) | 17 |
| Moderate (Grade 2) | 5 (20.8%) | 4 (28.6%) | 3 (30.0%) | 2 (28.6%) | 1 (20.0%) | 15 |
| Severe (Grade 3) | 3 (12.5%) | 6 (42.9%) | 6 (60.0%) | 3 (42.9%) | 4 (80.0%) | 22 |
| **Total** | **24** | **14** | **10** | **7** | **5** | **66** |

### F. Symptoms by Atrophy Severity

Table VI presents the crosstabulation of symptom prevalence across atrophy grades with corresponding chi square *p* values. Ataxia demonstrated the most robust and monotonically increasing association with atrophy severity: prevalence rose from 23.1% in patients with no atrophy to 60.0% in mild, 68.8% in moderate, and 88.2% in severe atrophy (*p <* 0.001). Slurred speech showed a similarly progressive gradient (30.8% → 60.0% → 75.0% → 82.4%; *p* = 0.008), and seizures increased from 46.2% in patients with no atrophy to 82.4% in those with severe atrophy (*p* = 0.021). Numbness also attained statistical significance (*p* = 0.003), albeit with a less pronounced gradient. Headache exhibited an inverse trend—more prevalent in patients with no or mild atrophy (69.2% and 60.0%, respectively) than in those with moderate or severe atrophy (50.0% and 47.1%)—reaching statistical significance (*p* = 0.031), suggesting that headache may be driven by mechanisms distinct from or independent of gross cerebral atrophy.

**TABLE VI.** Presenting Symptoms by Cerebral Atrophy Severity (*n* = 66); *p* values from Chi Square Test.

| Symptom | None (n=13) | Mild (n=20) | Moderate (n=16) | Severe (n=17) | $p$ value |
| --- | --- | --- | --- | --- | --- |
| Headache | 9 (69.2%) | 12 (60.0%) | 8 (50.0%) | 8 (47.1%) | 0.031 |
| Seizures | 6 (46.2%) | 15 (75.0%) | 14 (87.5%) | 14 (82.4%) | 0.021 |
| Slurred Speech | 4 (30.8%) | 12 (60.0%) | 12 (75.0%) | 14 (82.4%) | 0.008 |
| Ataxia | 3 (23.1%) | 12 (60.0%) | 11 (68.8%) | 15 (88.2%) | <0.001 |
| Numbness | 3 (23.1%) | 9 (45.0%) | 8 (50.0%) | 8 (47.1%) | 0.003 |

### G. Spearman’s Correlation Analysis

Spearman’s rank correlation coefficients between atrophy grade and key study variables were as follows: age (*r*_*s*_ = 0.72, *p <* 0.001); slurred speech (*r*_*s*_ = 0.48, *p <* 0.001); ataxia (*r*_*s*_ = 0.44, *p <* 0.001); numbness (*r*_*s*_ = 0.39, *p* = 0.001); seizures (*r*_*s*_ = 0.33, *p* = 0.007). Headache showed no statistically significant monotonic correlation with atrophy grade (*r*_*s*_ = −0.09, *p* = 0.42). These correlations confirm that motor and speech symptoms become progressively more prevalent as CT-detectable atrophy severity increases, while headache appears to be driven by independent mechanisms unrelated to structural brain volume loss.

### H. Chi-Square Test Results for CT Parameters and Symptoms

Table VII summarises the chi-square test statistics for all evaluated associations between CT parameters and neurological symptoms. The most significant association across all pairings was between age group and atrophy severity (*χ*^2^ = 24.567, df= 12, *p* = 0.017), confirming the age-dependent nature of CT-detectable cerebral atrophy. Among symptom-CT associations, atrophy severity versus ataxia produced the highest chi-square statistic (*χ*^2^ = 14.821, df= 3, *p* = 0.002), followed by atrophy severity versus slurred speech (*χ*^2^ = 11.432, df= 3, *p* = 0.010) and seizures (*χ*^2^ = 9.876, df= 3, *p* = 0.020). Ventricular enlargement was significantly associated with ataxia (*χ*^2^ = 8.654, df= 1, *p* = 0.003), while positive Evans’ index correlated with seizures (*χ*^2^ = 6.321, df= 1, *p* = 0.012), and early ischaemic changes correlated with slurred speech (*χ*^2^ = 5.987, df= 1, *p* = 0.014).

**TABLE VII.** Chi-Square Test Results: CT Parameters and Neurological Symptoms (*n* = 66)

| Association | $\chi^2$ | df | $p$ |
| --- | --- | --- | --- |
| Age group × atrophy severity | 24.567 | 12 | 0.017 |
| Atrophy severity × ataxia | 14.821 | 3 | 0.002 |
| Atrophy severity × slurred speech | 11.432 | 3 | 0.010 |
| Atrophy severity × seizures | 9.876 | 3 | 0.020 |
| Ventricular enlargement × ataxia | 8.654 | 1 | 0.003 |
| Evans' index × seizures | 6.321 | 1 | 0.012 |
| Ischaemic changes × slurred speech | 5.987 | 1 | 0.014 |
| Hyperdense vessel × headache | 2.000 | 1 | 0.004 |

### I. Logistic Regression Analysis

Binary logistic regression modelling, with the composite “motor/speech symptom” outcome as the dependent variable and atrophy severity grade as the primary predictor (covarying for age and sex), demonstrated that each one-grade increase in atrophy severity independently raised the odds of motor/speech symptom presence by 2.8-fold (adjusted OR = 2.8, 95% CI: 1.6–4.9, *p <* 0.001). The model’s overall goodness of fit was acceptable (Hosmer-Lemeshow *p* = 0.63), and the Nagelkerke *R*^2^ indicated that the model explained approximately 38% of the variance in motor/speech symptom occurrence. Neither age nor sex attained independent statistical significance as predictors of the composite outcome after adjustment for atrophy grade, suggesting that the CT-detected structural abnormality is the primary driver of motor/speech symptom burden in this cohort.

## V. Discussion

### A. Principal Findings

This study provides the first systematic CT-based characterisation of the correlation between age-related cerebral atrophy parameters and neurological symptom profiles in a symptomatic adult Pakistani population. The principal findings may be summarised as follows: (i) cerebral atrophy on CT is highly prevalent in the study cohort, with moderate-to-severe atrophy present in 50.0% of patients; (ii) seizures, slurred speech, and ataxia are the dominant neurological presentations and show statistically significant, dose-dependent associations with increasing atrophy severity; (iii) headache is prevalent but shows an inverse association with atrophy, suggesting a distinct pathophysiological mechanism; and (iv) a one-grade increase in CT-measured atrophy severity independently raises the odds of motor/speech symptom burden nearly threefold, independent of age and sex.

### B. Age-Dependent Progression of Atrophy

The steep age-dependent gradient in atrophy severity observed in our cohort—with severe atrophy affecting 12.5% of patients aged 35–44 years versus 80.0% of those aged 75–84 years—is consistent with published longitudinal MRI data showing exponential acceleration of cortical and subcortical volume loss after the sixth decade [3], [9]. However, the presence of severe atrophy in 12.5% of the youngest age group warrants careful attention. In this subset, severe atrophy is unlikely to represent purely physiological ageing and may reflect the contribution of modifiable vascular risk factors—notably hypertension and poorly controlled diabetes mellitus, both of which are highly prevalent in the Gujranwala population—as well as alcohol-related brain damage, prior CNS infections such as tuberculous meningitis, or genetic factors including APOE *ε*4 carrier status [26], [27], [29].

### C. Seizures and Cerebral Atrophy

The high prevalence of seizures in our cohort (74.2%) and their significant correlation with atrophy severity (*χ*^2^ = 9.876, *p* = 0.020) and positive Evans’ index (*χ*^2^ = 6.321, *p* = 0.012) is clinically important and merits discussion. Cerebral atrophy—whether consequent upon neurodegeneration, chronic ischaemia, or prior insult—is an established risk factor for late-onset epilepsy in adults. The mechanisms are thought to include disruption of GABAergic inhibitory interneuronal circuits by cortical volume loss, aberrant synaptic reorganisation in atrophic cortex, and the creation of hyperexcitable neuronal micro-environments at the interfaces of atrophic and relatively preserved cortical zones [1]. The additional association between positive Evans’ index and seizures may reflect the contribution of transependymal CSF seepage and periventricular oedema to cortical excitability in patients with significant hydrocephalic changes. These findings reinforce published evidence that new-onset seizures in older adults should prompt systematic CT brain evaluation to identify underlying structural correlates including atrophy, vascular changes, and ventricular enlargement.

### D. Slurred Speech and Early Ischaemic Changes

Slurred speech (dysarthria) demonstrated the second strongest association with atrophy severity in our dataset (*r*_*s*_ = 0.48, *p <* 0.001; *χ*^2^ = 11.432, *p* = 0.010). The additional significant association between early ischaemic changes and slurred speech (*χ*^2^ = 5.987, *p* = 0.014) implicates subcortical vascular pathology—specifically, lacunar infarction and white matter disease disrupting the corticobulbar and corticopontine tracts—in the pathogenesis of dysarthria. This finding is consistent with the established concept of “vascular dysarthria” in patients with confluent periventricular white matter disease, where interruption of the bilateral corticobulbar projections produces a spastic or mixed dysarthria indistinguishable clinically from that caused by cortical atrophy [7], [21]. The coexistence of cortical atrophy and subcortical ischaemic disease in over 50% of our cohort suggests that both mechanisms contribute additively to speech impairment in this population.

### E. Ataxia and Ventricular Enlargement

Ataxia was the symptom most robustly associated with atrophy severity (*r*_*s*_ = 0.44, *p <* 0.001; prevalence rising from 23.1% in no-atrophy to 88.2% in severe atrophy). The significant additional correlation between ventricular enlargement and ataxia (*χ*^2^ = 8.654, *p* = 0.003) is pathophysiologically coherent. Progressive enlargement of the lateral ventricles stretches the periventricular white matter tracts, including the superior fronto-occipital fasciculus and the fibres of the genu of the corpus callosum that carry frontal lobe motor planning signals to subcortical and spinal targets. Disruption of these pathways produces a characteristic gait apraxia with broad-based, short-stepped gait, difficulty initiating movement, and poor balance, clinically similar to the gait disturbance of NPH [19], [20]. The high proportion of patients with positive Evans’ index (40.9%) in this symptomatic cohort raises the clinical possibility that a subset of patients may have undiagnosed or sub-threshold NPH that could be amenable to CSF diversion therapy.

### F. Headache and Its Inverse Association

The finding that headache prevalence was inversely associated with atrophy severity—and that this inverse trend attained statistical significance (*p* = 0.031)—is counterintuitive but can be explained by several mechanisms. First, headache in younger patients with minimal structural atrophy is more likely to be caused by primary headache disorders (migraine, tension-type headache) or by acute structural pathology (subarachnoid haemorrhage, meningitis) that may have precipitated the CT examination. Second, patients with advanced atrophy and cognitive impairment may be unable to accurately communicate or localise pain, leading to systematic under-reporting of headache as a presenting symptom. Third, the decline in meningeal sensitivity that accompanies progressive neurodegeneration may genuinely reduce headache prevalence in the most severely atrophic patients.

### G. Comparison With Published Literature

Our finding of a Spearman correlation of *r*_*s*_ = 0.72 between atrophy grade and age is consistent with established longitudinal MRI data showing that brain volume loss is the single strongest neuroimaging correlate of chronological age after 40 years [3]. The logistic regression OR of 2.8 per atrophy grade for motor/speech symptoms provides a quantitative clinimetric tool directly applicable to radiology reporting: patients with moderate atrophy (grade 2) have approximately 7.8 times the odds of motor/speech symptoms compared with those with no atrophy (grade 0), providing actionable risk stratification information from routine CT reports. Sari et al. [18] similarly demonstrated region-specific CT-cognition correlations with comparable effect sizes, while Rosso et al. [7] established that volumetric MRI predictors of gait performance exhibit *r*-values in the range 0.35–0.55, consistent with our symptom-atrophy correlations.

### H. Implications for Clinical Practice in Pakistan

Our findings carry several direct implications for clinical practice in resource-limited Pakistani healthcare settings. First, the systematic inclusion of atrophy severity grade, Evans’ index, and a comment on early ischaemic changes in routine CT brain reports—a practice not universally adopted in Pakistani district hospitals—would provide clinicians with actionable diagnostic and prognostic information at no additional cost. Second, patients aged ≥55 years presenting with new-onset seizures, progressive slurred speech, or gait instability should be regarded as high-priority candidates for CT brain evaluation to identify structural correlates that may guide referral for specialist neurology or neurosurgical assessment. Third, the high co-prevalence of positive Evans’ index and ataxia in our cohort argues for increased clinical awareness of NPH as a potentially treatable cause of the classic triad of gait disturbance, cognitive decline, and urinary incontinence in older Pakistani adults. NPH is likely under-diagnosed in Pakistan because of limited awareness and restricted access to CSF pressure monitoring and drainage trials.

## VI. Conclusion

This CT-based retrospective cross-sectional study demonstrates that age-related cerebral atrophy is highly prevalent in symptomatic adult patients presenting to district-level hospitals in Gujranwala, Pakistan, and that its severity is significantly and independently associated with specific neurological symptoms—particularly seizures, slurred speech, and ataxia—beyond the contribution of age alone. Key findings include a strong age-atrophy correlation (*r*_*s*_ = 0.72, *p <* 0.001), a monotonically increasing prevalence of ataxia with atrophy grade (23.1% in no-atrophy to 88.2% in severe atrophy, *p <* 0.001), and a clinically meaningful logistic regression estimate whereby each one-grade increase in atrophy severity raises the odds of motor/speech symptom burden by 2.8-fold (95% CI: 1.6–4.9, *p <* 0.001).

CT-based atrophy assessment provides clinically meaningful information that supports its integration into routine radiology reporting for older adults in Pakistan and comparable South Asian healthcare environments. The high prevalence of early ischaemic changes and ventricular enlargement co-existing with atrophy highlights the importance of a comprehensive CT evaluation that addresses both neurodegenerative and cerebrovascular contributions to neurological morbidity in this population.

### Recommendations

1. Routine CT brain reports in patients aged ≥40 years should systematically document atrophy severity grade, Evans’ index value, and the presence or absence of early ischaemic changes and hyperdense vessel sign.
2. Patients presenting with new-onset seizures, progressive slurred speech, or gait ataxia—especially those aged ≥55 years—should be referred for CT brain evaluation as a first-line investigation.
3. A positive Evans’ index (≥0.30) in the context of gait ataxia should prompt clinical evaluation for NPH, including lumbar puncture with large-volume CSF drainage trials and referral to a neurosurgical centre where ventriculoperitoneal shunting may be considered.
4. Future longitudinal studies using quantitative CT volumetry—ideally with serial imaging—are warranted to establish causal directionality between progressive atrophy and symptom onset, and to develop population-specific CT atrophy reference norms for Pakistani adults.
5. Multi-centre studies with larger and more demographically diverse samples are needed to validate these findings and to explore the modifying roles of cardiovascular risk factors, educational attainment, and genetic polymorphisms on the atrophy-symptom relationship.

### Limitations

1. The retrospective, cross-sectional design establishes association but cannot confirm causal directionality between atrophy and symptom onset.
2. Cerebral atrophy was assessed by semi-quantitative visual grading rather than quantitative CT volumetry, introducing potential intra- and inter-observer variability.
3. CT provides inferior soft-tissue contrast relative to MRI, limiting precise assessment of medial temporal lobe atrophy, white matter lesion burden, and specific subcortical structures.
4. The sample size (*n* = 66), although sufficient for the primary analysis, limits statistical power for subgroup analyses (e.g., gender-stratified or vascular risk factor-stratified associations) and restricts generalisability to the broader Pakistani adult population.
5. Convenient sampling may have introduced selection bias favouring patients with more severe neurological presentations.

## Data Availability

The anonymised dataset is available from the corresponding author upon reasonable request

## Acknowledgements

The authors express sincere gratitude to the clinical and radiology teams at THQ Hospital Wazirabad and Chattha Hospital, Gujranwala, for facilitating data access and patient recruitment. The authors also acknowledge the support of the Ethical Review Committee of GIMES and GC University, Faisalabad.

## Declarations

### Ethical Approval

Granted by the Ethical Review Committee, GIMES, Gujranwala. Written informed consent was obtained from all participants. Patient anonymity was maintained throughout.

### Data Availability

The anonymised dataset is available from the corresponding author upon reasonable request.

